# Shared genetic basis informs the roles of polyunsaturated fatty acids in brain disorders

**DOI:** 10.1101/2023.10.03.23296500

**Authors:** Huifang Xu, Yitang Sun, Michael Francis, Claire F. Cheng, Nitya T.R. Modulla, J. Thomas Brenna, Charleston W. K. Chiang, Kaixiong Ye

## Abstract

The neural tissue is rich in polyunsaturated fatty acids (PUFAs), components that are indispensable for the proper functioning of neurons, such as neurotransmission. PUFA nutritional deficiency and imbalance have been linked to a variety of chronic brain disorders, including major depressive disorder (MDD), anxiety, and anorexia. However, the effects of PUFAs on brain disorders remain inconclusive, and the extent of their shared genetic determinants is largely unknown. Here, we used genome-wide association summary statistics to systematically examine the shared genetic basis between six phenotypes of circulating PUFAs (N = 114,999) and 20 brain disorders (N = 9,725-762,917), infer their potential causal relationships, identify colocalized regions, and pinpoint shared genetic variants. Genetic correlation and polygenic overlap analyses revealed a widespread shared genetic basis for 77 trait pairs between six PUFA phenotypes and 16 brain disorders. Two-sample Mendelian randomization analysis indicated potential causal relationships for 16 pairs of PUFAs and brain disorders, including alcohol consumption, bipolar disorder (BIP), and MDD. Colocalization analysis identified 40 shared loci (13 unique) among six PUFAs and ten brain disorders. Twenty-two unique variants were statistically inferred as candidate shared causal variants, including rs1260326 (*GCKR*), rs174564 (*FADS2*) and rs4818766 (*ADARB1*). These findings reveal a widespread shared genetic basis between PUFAs and brain disorders, pinpoint specific shared variants, and provide support for the potential effects of PUFAs on certain brain disorders, especially MDD, BIP, and alcohol consumption.

## Introduction

Disorders of the brain contribute significantly to the global disease burden [1, 2]. For example, in 2019, more than 970 million individuals suffered from 12 mental disorders, ranging from 13.6 million for eating disorders to 301.4 million for anxiety disorders [2]. These disorders encompass a wide range of psychiatric and neurological symptoms, including cognitive impairment, emotional dysregulation, and behavioral disturbances, all of which profoundly disrupt the life of the patients, and can in severe cases lead to suicide [3]. Effective prevention and treatment of brain disorders are of utmost importance in improving clinical symptoms and overall quality of life. One promising and emerging therapeutic approach is nutritional medicine [4], which seeks to prevent the onset of brain disorders or alleviate their clinical manifestations by implementing specific nutritional interventions [4, 5].

Brain structural lipids are rich in long-chain omega-3 and omega-6 polyunsaturated fatty acids (PUFAs) [6]. Dietary deficiency of omega-3 PUFAs leads to global deficits in neural function in experimental animals [7]. In humans, PUFAs and particularly omega-3s, have been suggested to have protective and therapeutic effects on brain disorders because they regulate physiological processes such as neuroinflammation, neurotransmission, and neuron survival [4, 6, 8]. Omega-3 supplementation has shown promising results in reducing clinical symptoms associated with a range of brain conditions, including MDD [9, 10], anxiety disorders [11], schizophrenia [12, 13], attention-deficit/hyperactivity disorder (ADHD) [14], autism spectrum disorder [15] and Alzheimer’s disease (ALZ) [16]. However, several randomized controlled trials reported small or no effects of PUFAs on schizophrenia [5], depression [17], ALZ [18] and psychosis [19, 20]. Consequently, the overall impact of PUFAs on human brain disorders remains inconclusive, necessitating further investigation to establish their therapeutic potential.

While observational associations are commonly confounded by unknown or unmeasured factors [21], exploring the shared genetic basis between PUFAs and brain disorders offers valuable insights into their shared biological pathways and potential causal relationships [22]. Previous studies have leveraged genetic information to investigate the connections between PUFAs and brain disorders (**Supplementary Table S1**), such as the application of Mendelian randomization (MR) to statistically infer causal relationships. For instance, a recent MR study suggested that decreased docosahexaenoic acid (DHA) and increased omega-6 to omega-3 ratio have causal links with MDD, and it further identified the fatty acid desaturase (*FADS*) gene cluster as a common genetic signal [23]. In an experimental study, mice with *Fads1/2* genes knockout were used to simulate the effect of BIP risk allele on *Fads1/2* activity, revealing significant changes in lipid profile and behavioral alterations [24]. However, current genetic studies primarily concentrate on specific brain disorders (e.g., MDD [23, 25], SCZ [26, 27], and BIP [24, 28]) or a limited number of genes, such as *FADS* [23, 24, 26, 28] and *ELOVL2/5* [26]. Therefore, it is necessary to explore the broader genomic landscape to ascertain additional genetic determinants that underlie the connection between PUFAs and brain disorders.

Our study aims to systematically explore the shared genetic basis between the levels of circulating PUFAs (cPUFAs) and brain disorders, infer their potential causal relationships, identify shared genomic regions, and pinpoint specific shared genetic variants. We performed four major analyses using genome-wide association study (GWAS) summary statistics for six cPUFA phenotypes (N = 114,999) and 20 brain disorders (N = 9,725-762,917). First, we estimated genetic correlation, and second, quantified the number of shared genetic variants, between cPUFA phenotypes and brain disorders. Third, we performed MR analysis to statistically infer causal associations between cPUFAs and brain disorders. Lastly, we conducted colocalization analysis and statistical fine-mapping to identify colocalized regions and pinpoint putative shared causal variants. Collectively, our study characterizes the shared genetic basis and informs the relationships between cPUFAs and brain disorders.

## Methods

### GWAS summary statistics and preprocessing

Six cPUFA phenotypes and 20 brain disorders were included in the study (**Supplementary Figure S1; Supplementary Table S2**). The six cPUFA traits were the relative percentages of total PUFAs, omega-3, omega-6, DHA, and linoleic acid (LA) in total fatty acids, and the omega-6 to omega-3 ratio. They are abbreviated as PUFA%, omega-3%, omega-6%, DHA%, LA%, and omega-6:omega-3, respectively. The 20 brain disorders included schizophrenia (SCZ) [29], MDD [30], BIP [31], obsessive-compulsive disorder (OCD) [32], anxiety disorders and factors (ANX) [33], post-traumatic stress disorder (PTSD) [34], anorexia nervosa (AN) [35], autism spectrum disorder (ASD) [36], Tourette syndrome (TS) [37], attention deficit-hyperactivity disorder (ADHD) [38], mood disorders (MOOD), insomnia (INS) [39], neuroticism (NE) [40], ALZ [41], opioid dependence (OD) [42], cannabis use disorder (CUD) [43], alcohol dependence (AD) [44], alcohol use disorder identification test total score (AUDIT_T), AUDIT focusing on alcohol consumption (AUDIT_C) and AUDIT focusing on the problematic consequences of drinking (AUDIT_P) [45].

Publicly available GWAS summary statistics of all cPUFAs and brain disorders were downloaded from IEU Open GWAS [46] and Psychiatric Genomic Consortium (PGC) [47]. GWAS summary statistics for insomnia [39] were downloaded from the Center for Nutrigenomics and Cognitive Research (CNCR, https://ctg.cncr.nl/software/summary_statistics). Multiple GWAS for each of seven brain disorders (i.e., SCZ, BIP, MDD, INS, ALZ, AN, ASD) were included for replication analysis (**Supplementary Table S2**). A total of 34 GWAS for brain disorders and 11 GWAS for cPUFAs were examined. Four GWAS were removed from our study for reasons including 1) no clear information indicating effect allele (n=2) [48, 49]; 2) incorrect data format (n=1) [50]; 3) the number of cases is less than 1000 (n=1) [51]. We focused on European ancestry to align ancestry across studies. Phenotypes associated with alcohol intake (AD, AUDIT_T, AUDIT_C, AUDIT_P) had pairwise genetic correlations less than 1 [45], and therefore were analyzed separately.

All GWAS summary statistics were harmonized to ensure data quality and consistency. Summary statistics of three GWAS from hg18 reference genome build were converted into hg19/GRCh37 genome build by Liftover [52]. MungeSumstats (v1.3.17) [53] was used to harmonize all GWAS summary statistics including: 1) uniformity in strand designation; 2) uniformity in SNP ID; 3) same effect allele; 4) effect size and standard error, or Z score are included; 5) hg19/GRCh37 reference genome build is used; 6) uniformity in the p-value format; 7) removal of InDels; 8) removal of SNPs with low genotype imputation quality (INFO < 0.3). After harmonization, a total of 10,568,861 SNPs for six cPUFAs and 1,147,602 to 14,124,455 SNPs for 20 brain disorders were included in the downstream analysis (**Supplementary Table S2**). For each trait, we mainly focused on the GWAS with the largest sample size, and the rest were presented in supplementary results.

### Estimation of SNP-based heritability (h^2^) and pairwise genetic correlation (r)

Linkage Disequilibrium Score regression (LDSC, v1.0.1) [54] was applied to estimate SNP-based heritability (h^2^) for each phenotype using GWAS summary statistics. For case-control traits, h^2^ was converted to the liability-scale by considering the disease prevalence and sample proportion (**Supplementary Table S2**). For quantitative traits, the observed-scale heritability was estimated.

Cross-trait LDSC [55] was used to compute pairwise genetic correlations (r_g_) using GWAS summary statistics between six cPUFAs and 20 brain disorders. Pre-computed reference panel LD score of European samples in the 1000 Genomes Project (1KGP) phase 3 [56] was downloaded from https://data.broadinstitute.org/alkesgroup/LDSCORE/eur_w_ld_chr.tar.bz2. SNP-based heritability and pairwise genetic correlation analyses were run using Hapmap3 SNPs with imputation INFO > 0.9 and minor allele frequency (MAF) > 1%. SNPs in the major histocompatibility complex (MHC) region were excluded. P-value cutoffs of 0.05, 0.001, and 0.05 divided by the number of tests (i.e., the Bonferroni-corrected threshold) were used to represent different levels of statistical significance. Genetic correlation coefficients and p-values were visualized using the R corrplot (v0.92) package [57].

### Estimation of polygenicity

To estimate the number of common variants that are associated with cPUFAs or brain disorders, a univariate Gaussian mixture model in MiXeR [58] was applied to the GWAS summary statistics. We restricted the univariate analysis to 19 brain disorder GWAS (corresponding to 15 unique phenotypes) with N > 46,000 to ensure statistical power. Five GWAS for ANX, OCD, TS, OD, AD had small sample sizes and were not included in the analysis. Pre-computed EUR reference panel LD score was used as in the LDSC analysis. To ensure compatibility with MiXeR, we utilized the munge_sumstats.py script provided by MiXeR to further process GWAS summary statistics. This step was necessary to meet the specific requirements of MiXeR, particularly addressing the sample imbalance in case-control phenotypes by utilizing the effective sample size 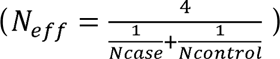. Additionally, we obtained information on allelic LD r^2^ correlations and allele frequency in the 1KGP European samples from the MiXeR GitHub repository. MiXeR provides a reference set of about 11 million SNPs, which is used to estimate the number of trait-associated variants that explain 90% of h^2^_SNP_.

### Quantification of polygenic overlap between cPUFAs and brain disorders

The MiXeR bivariate causal mixture model [59] was applied to quantify the number of variants that have nonzero effects on both traits (nc_12_). We performed cross-trait analyses to estimate polygenic overlap between cPUFAs and brain disorders, including six GWAS for six cPUFAs and 19 GWAS for 15 brain disorders. The bivariate analysis provides the proportion (π_12_), number (nc_12_), and correlation of effect size within the shared polygenic components (ρ_12_). We calculated Z-statistics using the formula Z /J/SE and visualized the effect sizes of all SNPs in pairs of GWAS summary statistics using the R hexbin (v1.28.2) package. We used R ComplexHeatmap (v2.14.0) [60] package to visualize the number of shared variants between cPUFAs and brain disorders.

### Mendelian randomization

MR is a method in genetic epidemiology that uses SNPs as genetic instruments to statistically infer causal associations between exposures and outcomes [61]. SNPs were identified as being significantly associated with each exposure at the genome-wide significance level (*P* < 5×10^-8^), and independent SNPs were derived using LD clumping (r^2^ < 0.001 within a 10,000 kb window). For the primary analysis, the potential causal effects were estimated using a multiplicative random-effect inverse weighted variance (IVW) model [62]. The MR-Egger method was applied to detect and correct for possible pleiotropy, while a p-value > 0.05 in its intercept test was used to rule out the presence of horizontal pleiotropic effects [63]. We also used weighted median and weighted mode approaches to explore the robustness of our findings in the presence of potential pleiotropy [64, 65]. As an additional sensitivity analysis against pleiotropy, the MR-PRESSO method was performed to evaluate overall horizontal pleiotropy and to re-calculate effect estimates after removing outlier SNPs [66]. A threshold of F-statistics > 10 indicates strong genetic instruments. Cochran Q-statistic was calculated to quantify the heterogeneity among SNPs [67, 68]. Scatter plots, forest plots, and leave-one-out plots were generated to visualize the effects of individual genetic instruments. To adjust for multiple testing, we utilized the false-discovery rate (FDR) approach [69]. All analyses were performed using the TwoSampleMR (v0.5.6) and MR-PRESSO (v1.0) packages in R [66, 70].

### Colocalization analysis

We assessed the colocalization of genetic associations across traits using HyPrColoc (v1.0) [71]. First, pairwise colocalization analyses were conducted for each pair of cPUFA and brain disorder. We further performed multi-trait colocalization analysis for all cPUFAs and brain disorders. We used the default prior probability that an SNP is associated with a single trait (*P* = 1 × 10^-4^) and a conditional prior probability that an SNP is associated with an additional trait given that it is already associated with another trait (*P*_c_ = 0.02). We defined a significant colocalized region as a posterior probability (PP) > 0.7. Regional association plots and colocalization probability plots were generated with gassocplot (v0.14.0) R package, and LD information was from 1KGP.

### Genome-wide statistical fine-mapping

To statistically infer genetic variants that are causally associated with cPUFAs and brain disorders, we performed genome-wide statistical fine-mapping with GWAS summary statistics using SuSiE (v0.12.27). We first defined significant loci for each GWAS. Each significant locus was determined as a region spanning 500kb above and below a top significant SNP (*P* < 5 × 10^-8^). After defining one locus, we eliminated this locus, searched for the most significant SNP in the remaining dataset, and defined the next locus. We iterated this process until no additional significant locus was found. Note that some loci overlap with each other, and the inclusion of LD information in the overlapped region is sometimes necessary for accurate fine-mapping. Since samples of all cPUFA phenotypes and some brain disorders were obtained from UK Biobank, we used LD matrices calculated based on 337,000 British-ancestry individuals in UK Biobank (UKBB-LD) [72]. All LD matrices files were downloaded from https://labs.icahn.mssm.edu/minervalab/resources/data-ark/ukbb_ld/. We extracted pairwise allelic LD correlations (r) for all SNPs in each defined locus. We summarized and reported 95% credible sets (CS) of all significant loci. Additionally, we identified SNPs within the CS of the cPUFAs and brain disorders dataset, which exhibited a posterior probability greater than 0.5 in at least one dataset.

### Functional annotation and gene set enrichment analysis

To assess the functional consequences of the potentially causal variants prioritized by HyPrColoc and SuSiE, we used the Ensembl Variant Effect Predictor (VEP) [73] for functional annotation, including their nearby genes, variant type and consequence, allele frequency in the 1KGP European sample, pathogenicity, and related phenotypes. Gene set enrichment analysis was conducted for candidate genes using the FUMA [74] GENE2FUNC module. GTEx v8 RNA-seq data [75] was used to examine tissue-specific expression patterns of candidate genes.

### Data and code availability

All GWAS summary statistics are publicly available as described above. All the code for this study was uploaded to GitHub for public access (https://github.com/Huifang-Xu/PUFA-BD).

## Results

### Widespread, moderate genetic correlations between cPUFAs and brain disorders

Genetic correlations (r_g_) between cPUFA phenotypes and brain disorders were estimated using LDSC. Consistent with previous studies, there were strong genetic correlations between cPUFAs [76] and between brain disorders [77-80] (**Supplementary Figure S2**). Widespread and moderate genetic correlations were observed between 16 brain disorders and six cPUFA relative measures, including PUFA%, omega-6%, LA%, omega-3%, DHA% and the omega-6:omega-3 ratio (**Figure 1A, Supplementary Figure S3 and Supplementary Table S3**). Out of the total 120 pairs, 77 pairs (64.2%) had *P* < 0.05 (average |r_g_| = 0.19), 43 pairs (35.8%) had *P* < 0.001 (average |r_g_| = 0.23), and 34 pairs (28.3%) showed significant genetic correlations after Bonferroni correction (*P* < 4.17 × 10^-4^, average |r_g_| = 0.22). Over 60% of the significant pairs (48/77 pairs with *P* < 0.05 and 22/34 pairs with *P* < 4.17 × 10^-4^) showed negative correlations between cPUFAs and brain disorders, suggesting that the shared genetic determinants are associated with higher cPUFA levels but with reduced risks of brain disorders, such as NE and PUFA% (**Figure 1C**).

**Figure 1:**
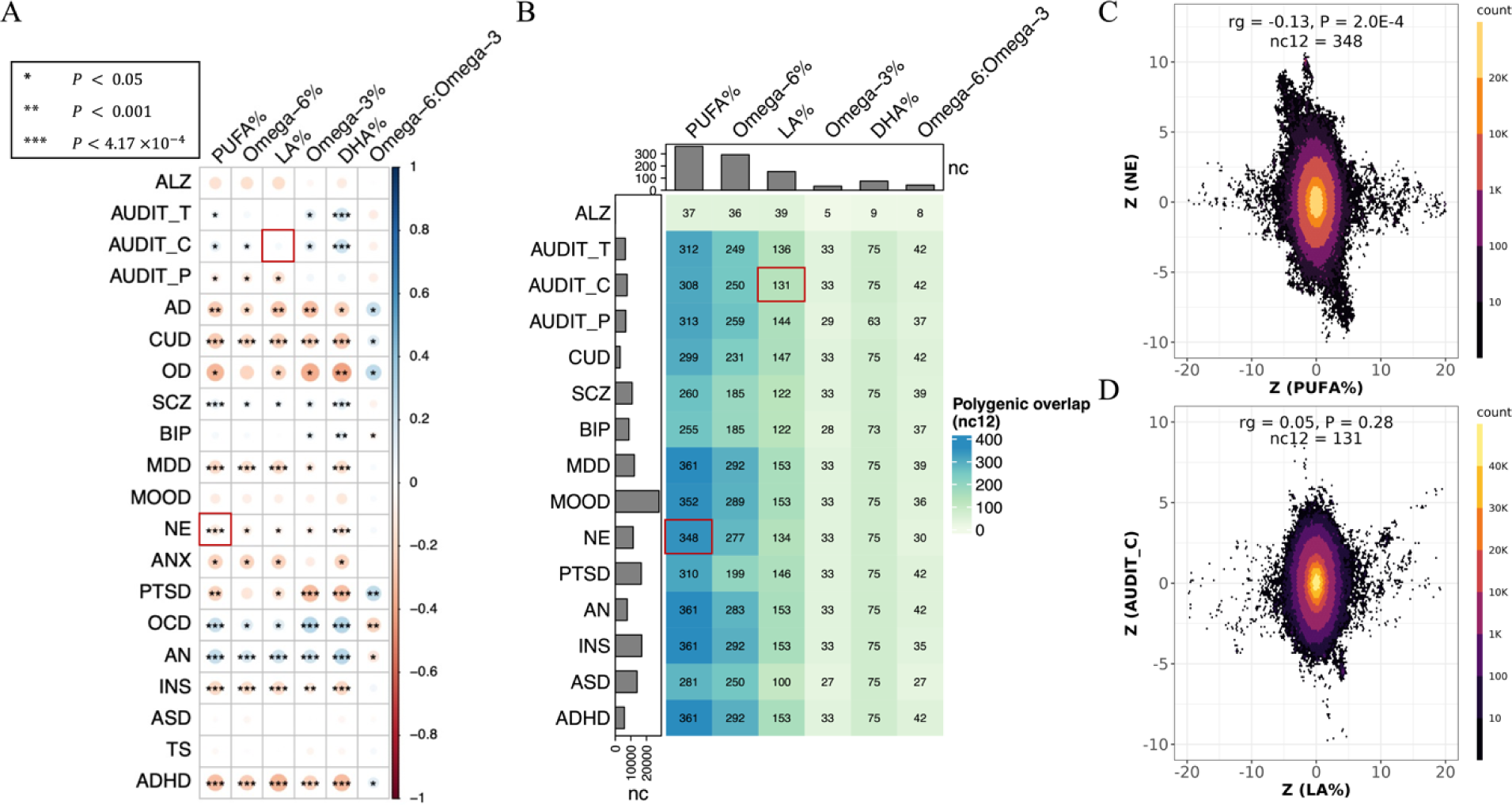
Widespread, moderate genetic basis shared between cPUFAs and brain disorders. **A)** Pairwise genetic correlations between six cPUFAs and 20 brain disorders. P-value cutoffs of 0.05, 0.001, 4.17×10^-4^ are used to represent increasing levels of statistical significance; colors are used to represent degree of genetic correlation (r_g_) between two traits. **B)** Pairwise polygenic overlaps between six cPUFAs and 15 brain disorders. The color and number of each box indicat the degree of polygenic overlap and number of causally associated SNPs shared between cPUFAs and brain disorders (nc_12_). Bar plots on the top and left indicate the number of cPUFAs- and brain disorders-associated variants, respectively, which explain 90% of SNP-based heritability. Two cPUFA-brain disorder pairs highlighted in the red boxes correspond to panels C) and D). **C)** Genetic effects of genome-wide SNPs on PUFA% (x axis) and NE (y axis). **D)** Genetic effects of genome-wide SNPs on LA% (x axis) and AUDIT_C (y axis). Each dot represents a genetic variant; colors indicate variant density.

PUFA%, omega-6%, omega-3%, LA%, and DHA% have significant negative correlation with the following brain disorders, including the three substance use disorders (OD: r_g_ = -0.23 ∼ - 0.40, *P* < 0.05; AD: r_g_ = -0.18 ∼ -0.30, *P* < 0.05; and CUD: r_g_ = -0.20 ∼ -0.27, *P* < 3 × 10^-4^), ADHD (r_g_ = -0.22 ∼ -0.33, *P* < 6.72 × 10^-6^), PTSD (r_g_ = -0.16 ∼ -0.32, *P* < 0.05), ANX (r_g_ = -0.22, *P* < 0.05), INS (r_g_ = -0.12 ∼ -0.20, *P* < 9 × 10^-4^), MDD (r_g_ = -0.10 ∼ -0.19, *P* < 0.05), and NE (r_g_ = -0.08 ∼ -0.14, *P* < 0.01; **Figure 1A** **and Supplementary Table S3**). In contrast, these cPUFA measures are positively correlated with two disorders with compulsive behaviors (OCD: r_g_ = 0.14 ∼ 0.30, *P* < 0.05; AN: r_g_ = 0.16 ∼ 0.27, *P* < 6.50 × 10^-5^). We did not observe any significant genetic correlations between any cPUFAs and ALZ, MOOD, ASD, or TS, suggesting that they share only a small proportion of common genetic components, or that the genetic components they share have mixed effects on the two traits. It can also be partially explained by insufficient statistical power due to small sample sizes of the GWAS of MOOD (N_case_ = 1,546) and TS (N_case_ = 4,819).

### Widespread, moderate polygenic overlap between cPUFAs and brain disorders

To quantify the polygenicity of and polygenic overlap between cPUFAs and brain disorders, we applied the MiXeR univariate and bivariate Gaussian mixture models, respectively, to their GWAS summary statistics. MiXeR statistically estimates the number of causal variants needed to explain 90% of the SNP heritability of a trait without explicitly identifying the specific variants. It also quantifies the number of shared causal variants between two traits (nc_12_), irrespective of their genetic correlation [59]. Five brain disorders (i.e., TS, OCD, ANX, OD and AD) were not included in this analysis due to insufficient sample sizes.

All pairs of cPUFAs and brain disorders were statistically inferred to share causal variants, although the degrees of sharing differ (**Figure 1B, Supplementary Figure S4 and Supplementary Table S4**). They ranged from five variants between omega-3% and ALZ to 361 between PUFA% and MDD. PUFA% shared the greatest number of common variants (nc_12_ = 37-361) with brain disorders, while omega-3% shared the least number of common variants (nc_12_ = 5-33). Consistent with the findings of genetic correlation, 10 brain disorders (MDD, CUD, AN, ADHD, NE, INS, SCZ, PTSD, AUDIT_C, and AUDIT_T) have strong polygenic overlaps with multiple cPUFAs. For instance, PUFA% and NE have a strong negative genetic correlation (r_g_ = - 0.13, *P* = 2.0 × 10^-4^) and a high level of polygenic overlap (nc_12_ = 348; **Figure 1C**), indicating that most of the common variants shared between PUFA% and NE have opposite effect signs. ALZ and cPUFAs share very low numbers of common variants. Interestingly, for some pairs of cPUFAs and brain disorders, we observed no significant genetic correlations; however, they have strong polygenic overlap, implying the presence of mixed effect directions among shared genetic variants. For example, LA% does not have significant genetic correlation with AUDIT_C (r_g_ = 0.05, *P* = 0.28), but they shared a moderate number of common variants (nc_12_ = 131). In addition, we found that the genetic variants they share had mixed effects on the two traits (**Figure 1D**), which explained why they had no significant genetic correlation but had strong polygenic overlap.

The numbers of shared variants between cPUFA levels and brain disorders are limited by the number of variants influencing cPUFAs. Compared with the strong polygenic overlap between different brain disorders (mean nc_12_ = 5,093; **Supplementary Figure S5 and Supplementary Table S5**), the average number of shared variants between cPUFAs is 76 (**Supplementary Figure S5**). We found that the number of shared variants is particularly limited by the number of variants underlying each specific cPUFA. The average number of common variants associated with cPUFA levels is 139, compared with 10,359 in brain disorders, a difference of two orders of magnitude (**Figure 1B; Supplementary Table S4**). Our polygenic overlap analysis revealed relatively simple genetic architecture of cPUFAs, high polygenicity of brain disorders, and widespread, moderate polygenic overlap between the two groups of traits.

### Statistical inference of causal associations between cPUFAs and brain disorders

To examine putative causal associations between six cPUFAs and 17 brain disorders, we conducted bidirectional MR analyses using GWAS summary statistics. Three brain disorders (AD, ALZ, and OD) were not included in the MR analysis due to the absence of effect sizes and standard errors in their GWAS summary statistics.

We identified nine pairs, for which genetically predicted cPUFAs were significantly (*P* < 0.05) associated with increased risks of brain disorders; and seven pairs, for which genetically predicted cPUFAs were associated with reduced risks of brain disorders (**Figure 2A-B** **and Supplementary Table S7**). Among the 16 significant pairs identified in the forward MR analysis, we did not detect any effect of brain disorders on cPUFA levels in our reverse MR analysis, except for the pair of PUFA%-MDD (**Supplementary Figure S6 and Supplementary Table S8**).

**Figure 2.**
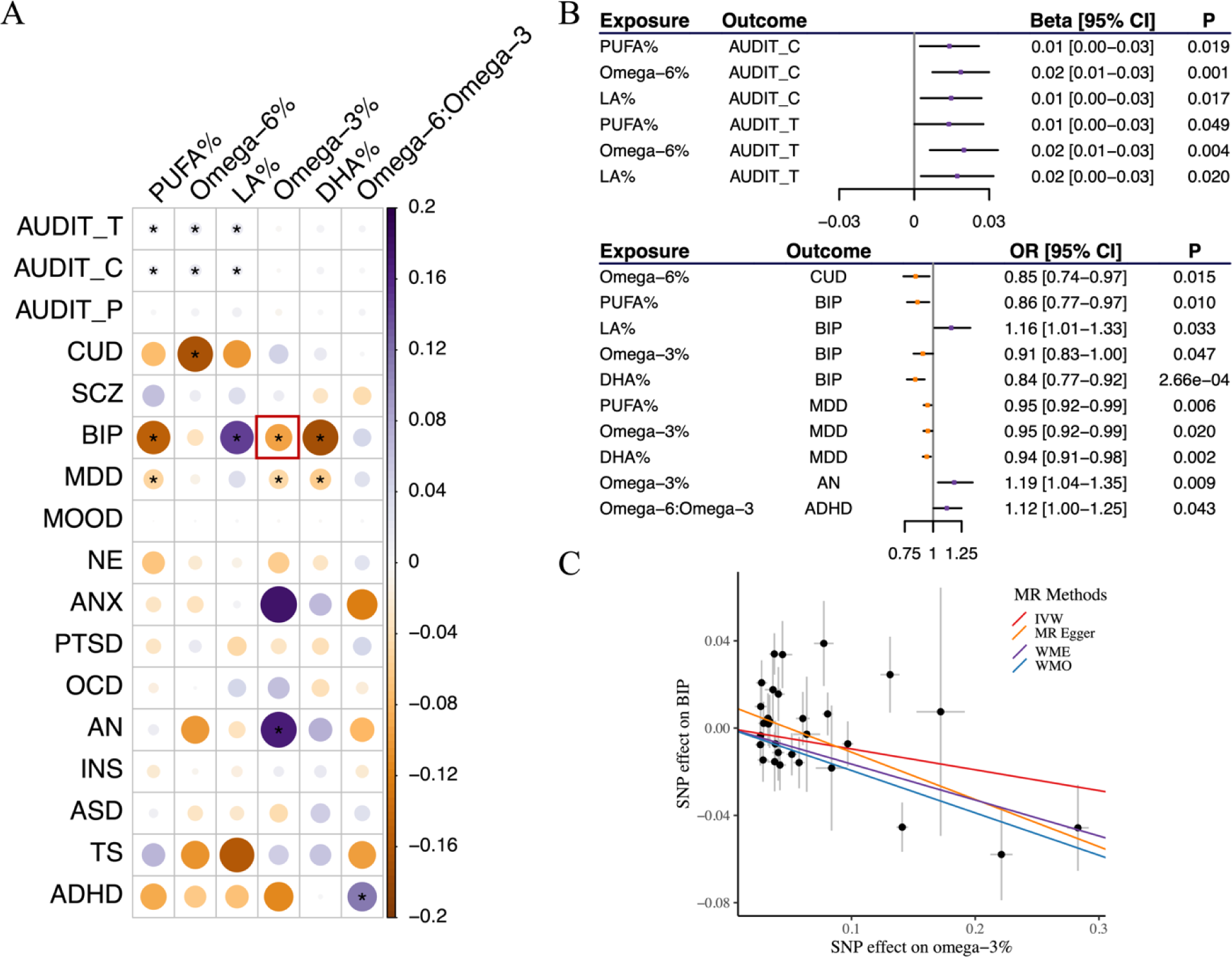
Statistical inference of causal relationship between cPUFAs and brain disorders. **A)** A heatmap summarizing the effects of six cPUFAs on 17 brain disorders. IVW p-value of 0.05 is used to represent statistical significance. Colors represent the effects (β_IVW_) of cPUFAs on brain disorders. The pair of omega-3% and BIP highlighted in the red box corresponds to panel C). **B)** MR results showing a significant association between cPUFAs and brain disorders. Beta and OR estimated using IVW method are used to represent the effects of cPUFAs on continuous and binary outcomes, respectively. **C)** MR estimated effects of omega-3% (x axis) on BIP (y axis). Effects estimated by the four models are shown by fitted lines; slopes of these line indicate the effect sizes.

Among the 16 significant pairs, nine pairs presented consistent and strong evidence for potential causal effects of cPUFAs on brain disorders when considering results from both genetic correlation and MR (**Figure 1A and 2A**). Four pairs (omega-6%-CUD, PUFA%-MDD, omega-3%-MDD, and DHA%-MDD) showed consistent negative associations, implying potential protective effects of these cPUFAs against CUD and MDD. In contrast, five pairs (PUFA%-AUDIT_C, omega-6%-AUDIT_C, PUFA%-AUDIT_T, omega-3%-AN, and omega-6:omega-3-ADHD) showed consistent positive associations, indicating that these cPUFAs might increase the risks of alcohol consumption, anorexia nervosa and ADHD.

Omega-3% were genetically predicted to be associated with a reduced risk of BIP. For a one standard deviation (SD) increase in genetically predicted omega-3%, the odds ratio (OR) for BIP was 0.91 (95% CI = [0.83, 1.00]) using the IVW method (**Figure 2C** **and Supplementary Table S7**). Although horizontal pleiotropy was detected in the intercept test (*P*_intercept_ = 0.043), the result remained significant after correcting for possible pleiotropy with the MR-Egger approach (OR = 0.81, 95% CI = [0.70, 0.93]). The finding was consistent across other MR methods. In the reverse MR, there was no evidence supporting a causal effect of BIP on omega-3% (β_IVW_ = -0.10, 95% CI = [-0.33, 0.13]) (**Supplementary Table S8**).

### Prioritization of colocalized loci and shared variants

To statistically prioritize genomic loci and infer causal variants responsible for both cPUFA levels and brain disorders, we conducted pairwise colocalization analysis and statistical fine-mapping. This analysis revealed 44 significant colocalized regions with a PP > 0.7 (**Figure 3A** **and Supplementary Table S9**). The 44 significant colocalized regions correspond to 13 unique regions. Furthermore, 22 unique SNPs were statistically inferred as potential causal variants shared between cPUFAs and brain disorders, indicating that more than one variant within these colocalized regions contribute to multiple trait pairs. Among the 22 unique SNPs, 14 were also included in 95% CSs defined by SuSiE (**Supplementary Table S9**). We also performed multi-trait colocalization analysis combining all cPUFAs and brain disorders. We identified four candidate shared SNPs (**Supplementary Table S9**).

**Figure 3.**
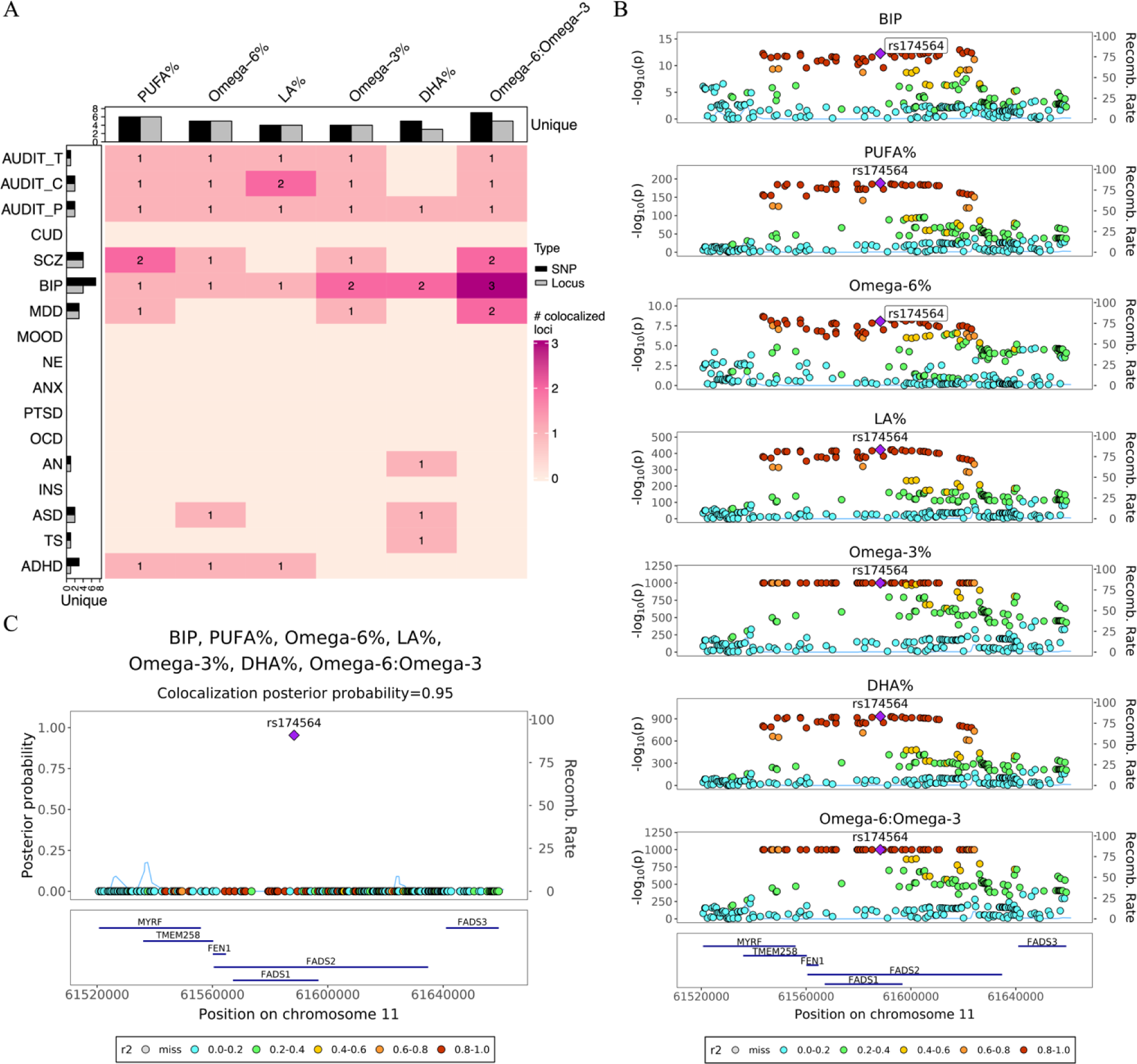
Colocalization analysis detects genomic loci shared between cPUFAs and brain disorders. **A)** A heatmap summarizing pairwise colocalization between six cPUFAs and 17 brain disorders. The color and number of each box indicate the number of significant colocalized regions between cPUFAs and brain disorders (PP > 0.7). Bar plots on the top and left indicate the numbers of unique colocalized SNPs (black) and loci (grey, PP > 0.7) for cPUFAs and brain disorders, respectively. **B)** Regional association plots of six cPUFA phenotypes and BIP in chr11:61,520,000-61,660,000. Variant positions are shown on x axis, -log_10_P on the left y axis, recombination rate on the right y axis; variant rs174564 is marked as the lead SNP; genes located in the region are shown in the bottom. LD r^2^ values are indicated by colors, and recombination rates by curves. **C)** Multi-trait colocalization analysis combing six cPUFA phenotypes and BIP identified a putative shared causal variant rs174564 (PP = 0.95). PP values are shown on y axis.

To gain insights into the functional implications of the identified colocalized SNPs, we annotated the nearest genes associated with the colocalized and fine-mapped SNPs using VEP (**Supplementary Table S10**). Additionally, we performed gene set enrichment analysis using the FUMA GENE2FUNC function [74]. This analysis revealed that the 36 prioritized genes are significantly enriched in biological pathways related to lipid metabolism (FDR adjusted *P* < 0.05), providing further support for their potential biological relevance in the context of cPUFA levels and brain disorders (**Supplementary Table S11**).

We highlight here one example that provides insights into the role of PUFAs on brain disorders. The example involves BIP and all six cPUFA measures (**Figure 3B and 3C**), all of which share a colocalized region at the *FADS* gene cluster (chr11:58,780,549-62,223,771). Within this region, three distinct shared SNPs (i.e., rs174564, rs174567, rs174528) were identified (**Supplementary Table S9**). To further investigate this region, we performed statistical fine-mapping analysis using SuSiE, which supported the presence of multiple causal variants for omega-3%, DHA%, PUFA% and omega-6:omega-3 (**Supplementary Figure S7**). This analysis provided additional evidence for the potential involvement of multiple causal variants within the *FADS* region in modulating the circulating levels of omega-3%, DHA%, PUFA%, and omega-6:omega-3.

We also performed a multi-trait colocalization analysis combining these six cPUFA phenotypes with BIP. The variant rs174564 (chr11:61588305A>G) had the highest PP of 0.95 (**Figure 3C**), suggesting that it is likely the shared causal variant between cPUFAs and BIP. The SNP is an intronic variant of the *FADS2* gene and is known to be associated with both cPUFA levels and BIP. The A allele of rs174564 was associated with an increased level of DHA% (β = 0.28, SE = 0.004, *P* < 1 × 10^-300^) and omega-3% (β = 0.39, SE = 0.004, *P* < 1 × 10^-300^), while with a reduced risk of BIP (OR = 0.93, 95% CI = [0.91, 0.95], *P* = 6.24 × 10^-13^). Furthermore, MR analysis also revealed a negative association between omega-3% and BIP (**Figure 2A-C**). Combining the results of MR and colocalization analysis, there is strong evidence supporting that omega-3% has a protective effect on bipolar disorder.

### Potential causal relationships informed by shared genetic basis

To advance our understanding of the potential causal relationship between cPUFAs and brain disorders, we compared and synthesized the findings across the multiple approaches of evaluating shared genetic basis. We designated strong evidence supporting a potential causal relationship when there are statistically significant and directionally consistent results in genetic correlation (*P* < 0.05), MR (*P* < 0.05), and colocalization (PP > 0.7). We did not include polygenic overlap due to its ubiquity among all six cPUFA phenotypes and brain disorders. We considered that there is suggestive evidence when there were statistically significant and directionally consistent results in genetic correlation (*P* < 0.05) and colocalization (PP > 0.7).

We identified four pairs with strong evidence supporting potential causal effects of the specific cPUFAs on the corresponding brain disorders (**Table 1**). For example, PUFA% is likely to have protective effect on MDD with support from the following evidence: 1) PUFA% showed a negative genetic correlation with MDD (r_g_ = -0.19, *P* = 7.14 × 10^-16^); 2) MR results suggest that higher PUFA% is associated with a reduced risk of MDD (OR = 0.95, 95% CI = [0.92, 0.99], *P* = 5.76 × 10^-3^); 3) Colocalization analysis identified a colocalized region at chr21q22.3 (chr21:46,177,105-47,492,226; PP = 0.83) and a potential shared causal variant rs4818766 (chr21:46635351A>G), which is an intronic variant of gene *ADARB1*. SNP rs4818766 is known to be associated with body fat distribution [81]. *ADARB1* is highly expressed in the brain and related to developmental and epileptic encephalopathy [82] and psychiatric disorders [83, 84]. In addition to PUFA%, our forward MR results also show that higher levels of omega-3% and DHA% were associated with a reduced risk of MDD, in line with a recent finding [23]. Interestingly, our forward and reverse MR both showed negative associations between PUFA% and MDD (forward MR: OR = 0.95, 95% CI = [0.92, 0.99], *P* = 5.76 × 10^-3^; reverse MR: OR = 0.91, 95% CI = [0.84, 0.99], *P* = 0.013), driven by different genetic variants (**Supplementary Figure S8**), supporting a potential bidirectional relationship.

**Table 1.**
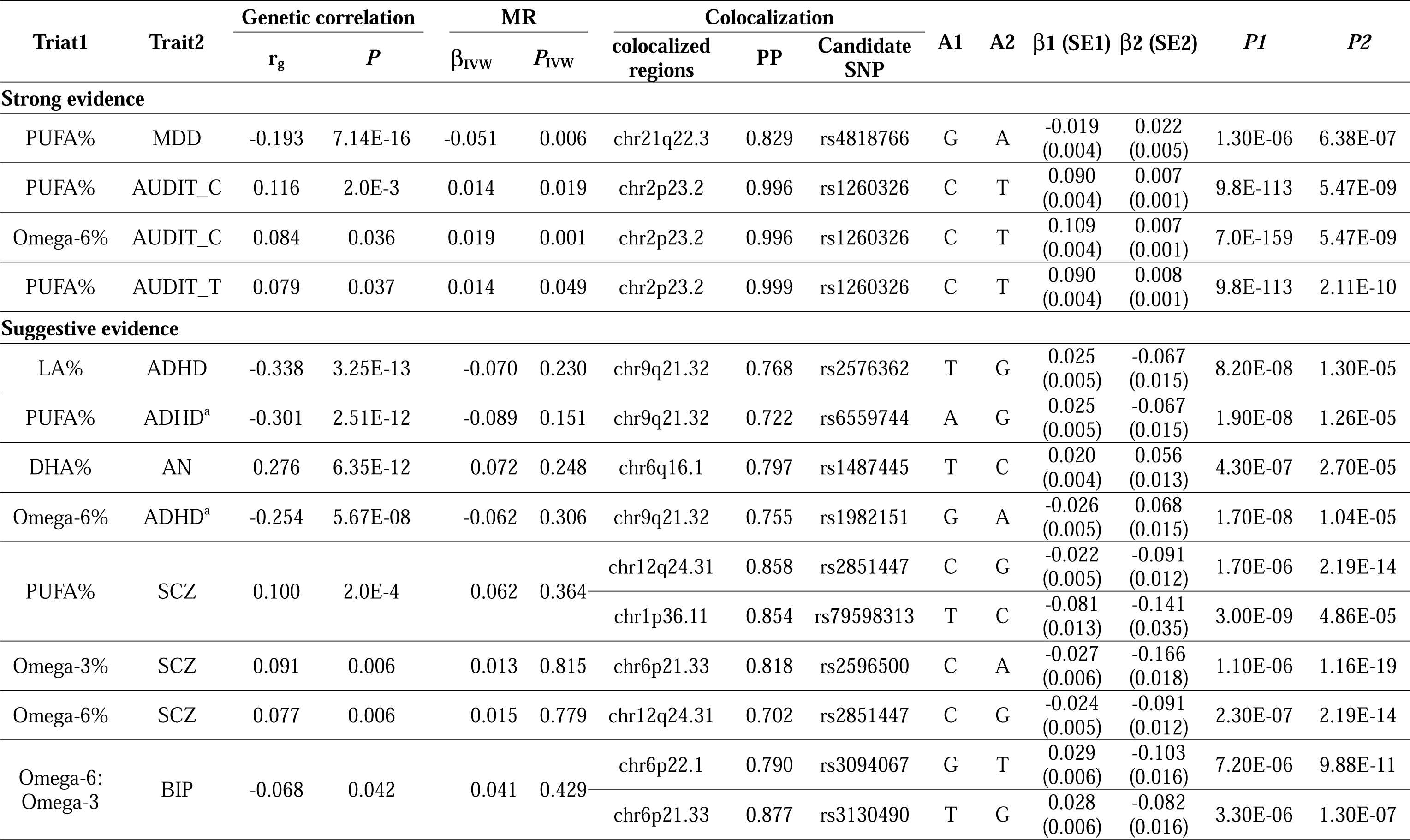
Evidence supporting the potential role of cPUFAs in brain disorders. Note: r_g_, genetic correlation; β_IVW_, estimated effects of trait 1 on trait 2 using IVW method; PP, posterior probability. A1, effect allele; A2, reference allele. β1, SE1 and β2, SE2 are genetic effects and standard errors of A1 on trait 1 and trait 2, respectively. *P1* and *P2* are *P* values for trait 1 and trait 2 extracted from GWAS summary statistics, respectively. _a_The reverse MR of the two PUFA-ADHD pairs showed significant results (*P*_IVW_ < 0.05).

We also found that lower omega-6% are related to lower alcohol consumption (**Table 1**). Both genetic correlation (r_g_ = 0.08, *P* = 0.036; **Figure 4A**) and forward MR results (β_IVW_ = 0.019, *P* = 0.001; **Figure 4B**) revealed a positive association between omega-6% and alcohol consumption. In our colocalization analysis (**Figure 4C and 4D**), we observed that genomic region 2p23.2-2p23.3 (chr2:26,894,985-28,598,777) exhibited colocalization signals among three alcohol-intake phenotypes (AUDIT_C, AUDIT_T, AUDIT_P) and five cPUFA phenotypes (omega-3%, omega-6%, LA%, PUFA% and omega-6:omega-3). Within this region, SNP rs1260326 (chr2:27730940T>C) was identified as a potential shared causal variant (PP = 0.99). Notably, the T allele of rs1260326 was associated with lower levels of omega-6% (β = -0.11, SE = 0.004, *P* = 7.0 × 10^-159^), LA% (β = -0.08, SE = 0.004, *P* = 1.20 × 10^-87^) and lower alcohol consumption (AUDIT_C: β = -0.007, SE = 0.001, *P* = 5.47 × 10^-9^; AUDIT_T: β = -0.008, SE = 0.001, *P* = 2.11 × 10^-10^; AUDIT_P: β = -0.005, SE = 0.001, *P* = 6.7 × 10^-7^). SNP rs1260326, a missense variant for gene *GCKR*, is known to be associated with alcohol intake [85], type 2 diabetes [86], liver diseases [87, 88] and lipid levels such as triglyceride and cholesterol [89]. Taken together evidence from genetic correlation, MR and colocalization analysis, our findings indicate that lower omega-6% may lower alcohol consumption.

**Figure 4.**
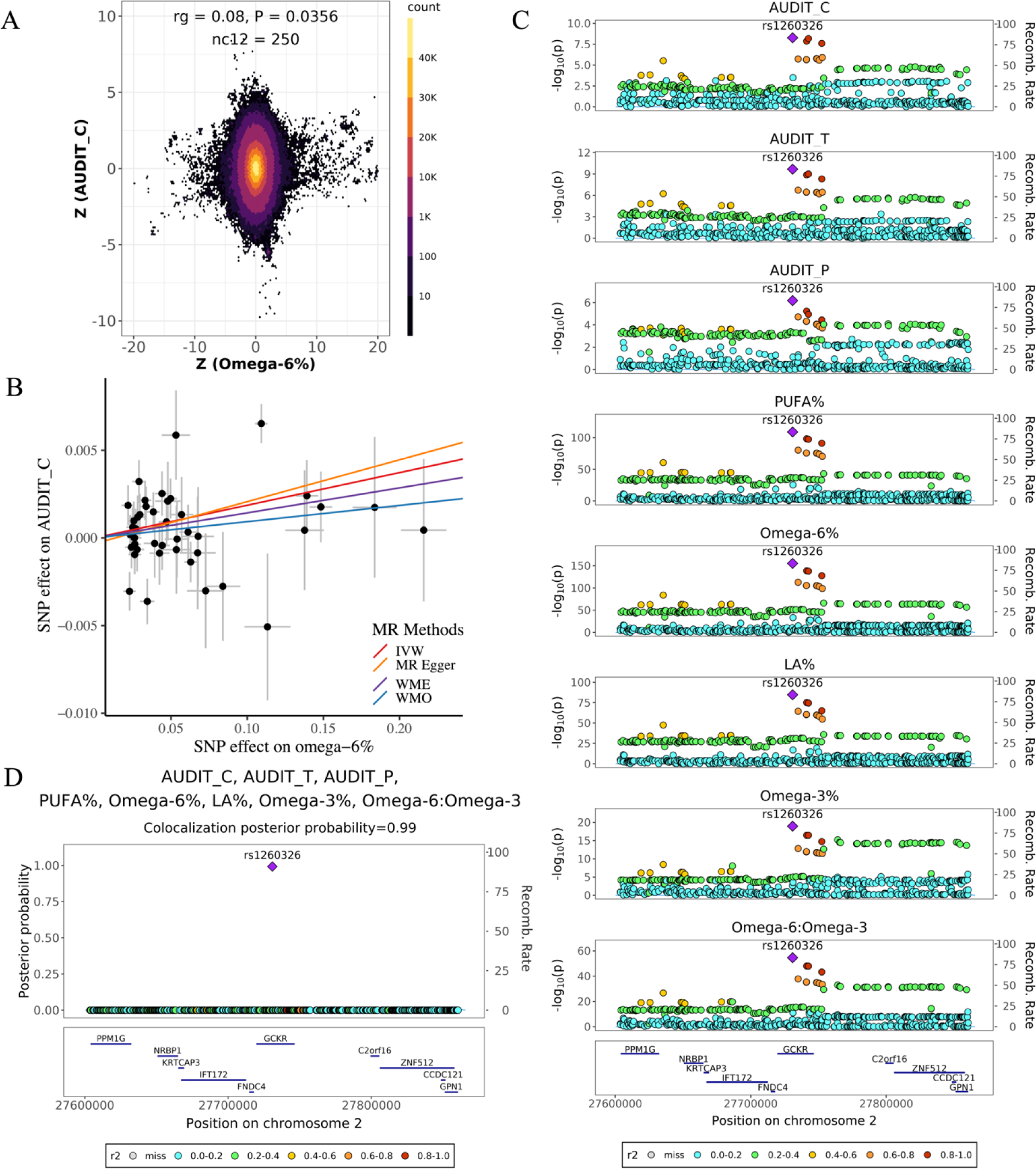
Evidence supporting the effect of omega-6% on alcohol consumption. **A)** Genetic effects of genome-wide SNPs on omega-6% (x axis) and AUDIT_C (y axis). Each dot represents a genetic variant; colors indicate the variant density. **B)** MR estimated effects of omega-6% (x axis) on AUDIT_C (y axis). Effects estimated by the four models are shown by fitted lines;slopes of these lines indicate the effect sizes. **C)** Regional association plots of five cPUFA phenotypes and three alcohol consumption phenotypes in chr2:276,000,000-27,900,000. Variant positions are shown on x axis, -log_10_P on the left y axis, recombination rate on the right y axis; variant rs1260326 is marked as the lead SNP; genes located in the region are shown in the bottom. LD r^2^ values are indicated by colors, and recombination rates by curves. **D)** Multi-trait colocalization analysis combing five cPUFA phenotypes and three alcohol consumption phenotypes identified a shared putative causal variant rs1260326 (PP = 0.99). PP values are shown on y axis.

We identified eight trait pairs that display suggestive evidence for a potential causal relationship. Interestingly, our genetic correlation analysis unveiled a negative correlation between ADHD and three cPUFA phenotypes: PUFA% (r_g_ = -0.3, *P* = 2.51 × 10^-12^), omega-6% (r_g_ = -0.25, *P* = 5.67 × 10^-8^), and DHA% (r_g_ = -0.32, *P* = 1.73 × 10^-10^). Further colocalization analysis identified a genomic locus chr9:85,440,801-86,938,196 shared among ADHD, PUFA%, omega-6% and LA% (PP > 0.7; **Table 1**). Our forward MR did not reveal significant associations between ADHD and the three cPUFAs. However, the reverse MR displayed significant negative associations (PUFA%: β_IVW_ = -0.07, *P* = 3.48 × 10^-3^; omega-6%: β_IVW_ = -0.05, *P* = 0.046; and DHA%: β_IVW_ = -0.08, *P* = 3.43 × 10^-3^; **Supplementary Table S8**), suggesting that the presence of ADHD might contribute to decreased circulating PUFA levels. These findings align with previous research indicating that individuals with ADHD generally exhibit lower omega-3 PUFA levels compared to the control group [14]. This ADHD example provides clues for further studies into the intricate relationship between ADHD and cPUFAs.

## Discussion

By leveraging GWAS summary statistics of six cPUFA phenotypes and 20 brain disorders, we revealed a widespread shared genetic basis between the two groups of traits. Our MR analysis found 16 pairs of cPUFAs and brain disorders that display potential causal associations. Further colocalization and fine-mapping analysis led to statistically inferred candidate shared causal variants, such as rs1260326 (*GCKR*), rs174564 (*FADS2*) and rs4818766 (*ADARB1*). We also identified cPUFA-brain disorder pairs with consistent results across various analysis approaches, emphasizing a prominent role of cPUFAs in brain disorders, especially MDD, BIP and alcohol consumption-related phenotypes. Our discoveries provide novel insights into the intricate relationships between cPUFAs and brain disorders, improving our knowledge in refining dietary strategies for prevention and intervention.

The protective effect of PUFA% on MDD is strongly supported by various methods with different model assumptions, including genetic correlation, MR and colocalization. We identified a putative shared variant rs4818766 and a candidate gene *ADARB1*. *ADARB1* encodes one of the enzymes involved in the adenosine-to-inosine (A-to-I) RNA editing process known as Adenosine Deaminases Acting on RNA (ADAR2) [90]. One of the leading hypotheses regarding the pathogenicity of MDD is the serotonin hypothesis, which suggests that depression may arise from abnormalities in neurotransmitters, particularly serotonin [90, 91]. ADAR2 could edit serotonin 2C receptor (5-HT_2c_-R) at the D site, which reduces G protein coupling and affinity for serotonin [90]. Notably, prior research has shown that ADAR2 knock-out and mutant mice lacking the deaminase activity of ADAR2 exhibit elevated body fat and reduced ability to utilize fatty acids [92, 93]. Animal studies have also demonstrated that supplementing PUFAs in rats leads to higher concentrations of serotonin in the brain [94]. Taken together, it is plausible that PUFAs reduce the risk of MDD by modulating the serotonin transportation through ADAR2 [95].

Our study also supports the protective effect of omega-3% on BIP. MR analysis showed that higher omega-3% are associated with a reduced risk of bipolar disorder. Further colocalization analysis identified a colocalized region where the *FADS1* and *FADS2* genes are located. Statistically inferred shared causal variant rs174564 is an intronic variant of the *FADS2* gene. SNPs in the *FADS1/2* region have been reported to be associated with circulating PUFA levels and the risk of bipolar disorder in different populations [28, 96, 97]. Significant changes in the lipid profiles of the plasma and brain, as well as behavioral changes (e.g., hyperactivity and hypoactivity episodes), were observed in heterozygous *Fads1/2* knockout mice [24]. Moreover, dietary DHA supplementation reduced depressive episodes in the mutant mice, supporting the protective role of omega-3% against BIP.

We show that lower levels of omega-6% are related to lower alcohol consumption. We statistically inferred a shared causal variant rs1260326 (gene: *GCKR*), which explains a colocalized association signal between omega-6% levels and alcohol consumption. *GCKR* encodes glucokinase regulatory protein that binds to glucokinase. Compared to the C allele of rs1260326, the T allele results in lower binding efficiency of glucokinase regulatory protein, leading to increased total fatty acids formation, liver fat and triglyceride accumulation [98]. In addition, the T allele is linked to a higher risk of liver diseases, including nonalcoholic fatty liver disease (NAFLD) and non-alcoholic steatohepatitis [99]. Lower serum levels of omega-6 fatty acids and LA were associated with a higher risk for NAFLD [100]. Taken together, it is possible that individuals with lower PUFA and omega-6 levels tend to have more liver problems (e.g., accumulation of liver fat, elevated levels of triglyceride, and alanine aminotransferase), and thus tend to drink less.

We note that disease status itself might influence cPUFA levels. Our reverse MR results revealed a significant negative association between ADHD and three cPUFAs (PUFA%, omega-6% and LA%), suggesting that altered cPUFA levels may be one of the metabolic consequences of ADHD. Further pairwise colocalization analysis identified a region chr9:85,440,801-86,938,196 colocalized among ADHD, PUFA%, omega-6% and LA% (**Table 1** and **Supplementary Table S9**). Three distinct SNPs (i.e., rs2576362, rs1982151, rs6559744) were statistically inferred as putative causal variants that explain the shared association signal. However, none of the identified SNP has strong enough evidence for causation (PP < 0.1), and further studies are needed to pinpoint shared causal variants and candidate genes in this region.

The discrepancy between the genetic correlation and MR results could be attributed to the differences in the sets of genetic variants analyzed in either approach and the existence of discordant pleiotropy across variants. It also reflects the limitations of different methods as well as the complex genetic architecture of brain disorders [101]. Taking omega-3% and BIP as an example, their positive genetic correlation suggests the presence of a substantial number of common variants that exert small yet consistent effects on both phenotypes (**Supplementary Figure S9)**. However, the negative association observed in MR and colocalization analysis is driven by the *FADS* locus that exhibited a relatively large effect but with opposite directions on the two phenotypes (**Supplementary Figure S8 and Supplementary Table S12)**. We highlight the need to understand the biological function of genetic variants in MR analysis, especially when the trait of interest has complicated genetic architecture [101, 102].

In our analysis, we focused on relative measures of cPUFAs. We found limited genetic correlations or polygenic overlaps between the absolute measures of cPUFAs and brain disorders (**Supplementary Figure S3-S4 and Table S3-S4**). It is important to note that absolute and relative measures of cPUFAs offer distinct perspectives on fatty acid metabolism. Relative measures are preferred in the majority of cases because PUFAs as well as saturated and monounsaturated fatty acids are metabolized by the same enzymes derived from common genes (e.g. *FADS1*/*FADS2*) [103]. They are also preferred because relative measures are more precise (lower analytical SDs) since they are all referenced to one another and not to exogenously-added internal standards. Absolute measures provide direct information about the quantities of cPUFAs which become important when any particular fatty acid may become limiting for a particular physiological requirement [104-106]. The different patterns with brain disorders are consistent with the limited genetic correlation between absolute and relative cPUFAs (Supplementary Figure S2). They likely reflect different aspects of lipid metabolism. Future studies are needed to discern the exact mechanisms.

Our study is not without limitations. First, using different GWAS summary statistics could lead to minor differences in the results since slightly different analytical strategies were applied (e.g., association methods, quality control criteria, and covariates). To address this issue, we analyzed multiple GWAS of the same phenotype to evaluate robustness of our discoveries. We observed consistent correlation patterns with different GWAS of the same traits (**Supplementary Figure S3 and Supplementary Table S3**). Second, five brain disorders (i.e., ANX, OCD, OD, AD, and TS) had relatively small sample sizes that did not meet the requirement of MiXeR and were therefore excluded from the estimation of polygenic overlap. Finally, our study focused only on the European population. Genetic adaptation and variation of fatty acid composition have been demonstrated in the Inuit, African, South Asian, East Asian, and European populations [23, 107-109]. Differences in prevalence [110, 111] and genetic risk factors [89] of psychiatric disorders were also demonstrated across ethnic groups. Therefore, expanding our research to other populations is necessary to gain a deeper understanding of the shared genetic basis and genetic determinants between cPUFAs and brain disorders across populations.

Our systemic genetic analysis of six cPUFA traits and 20 brain disorders uncovered a widespread shared genetic basis between the two groups. We pinpointed specific shared genetic variants and provided evidence supporting the potential effects of certain cPUFAs on specific brain disorders. Our findings provide new insights into the shared genetic architecture underlying these traits and have implications for interventions and dietary recommendations of PUFAs in the context of brain disorders.

## Supplemental information description

Figure S1. Flowchart of the study

Figure S2. Pairwise genetic correlations A) between brain disorders and B) between cPUFA phenotypes.

Figure S3. Pairwise genetic correlations between cPUFAs and brain disorders Figure S4. Pairwise polygenic overlaps between cPUFAs and brain disorders

Figure S5. Pairwise polygenic overlaps A) between brain disorders and B) between cPUFAs.

Figure S6. Reverse MR results

Figure S7. Statistical fine-mapping posterior inclusion probability plots in 11q12.1-11q12.3 for A) omega-3%, B) DHA%, C) PUFA% and D) omega-6:omega-3

Figure S8. Bidirectional association between PUFA% and MDD

Figure S9. Differences in the SNP sets in omega-3%-BIP genetic correlation and MR analysis

Table S1. Literature review on PUFA-brain disorders Mendelian randomization studies

Table S2. Dataset characteristics

Table S3. Pairwise genetic correlations between cPUFAs and brain disorders

Table S4. MiXeR univariate estimates and bivariate estimates between cPUFAs and brain disorders

Table S5. MiXeR bivariate estimates between brain disorders

Table S6. MiXeR bivariate estimates between cPUFAs

Table S7. Forward MR results inferring the causal effect of cPUFAs on brain disorders

Table S8. Reverse MR results inferring the causal effect of brain disorders on cPUFAs levels

Table S9. Pairwise colocalization analysis and fine-mapping results

Table S10. Functional annotation of colocalized and fine-mapped SNPs by VEP

Table S11. Gene-set enrichment analysis of 36 candidate genes using FUMA GENE2FUNC

Table S12. Genetic instruments included in the omega-3%-BIP MR analysis

## Data Availability

All GWAS summary statistics are publicly available as described in the Methods section. All the code for this study was uploaded to GitHub for public access (https://github.com/Huifang-Xu/PUFA-BD).

## Abbreviations

1KGP: 1000 Genomes Project
AD: Alcohol dependence
ADARB1: Adenosine Deaminase RNA Specific B1
ADHD: Attention deficit-hyperactivity disorder
ALZ: Alzheimer’s disease
AN: Anorexia nervosa
ANX: Anxiety disorders and factors
ASD: Autism spectrum disorder
AUDIT: Alcohol use disorder identification test
AUDIT_C: AUDIT focusing on alcohol consumption
AUDIT_P: AUDIT focusing on the problematic consequences of drinking
AUDIT_T: AUDIT total score
BIP: Bipolar disorder
cPUFA: Circulating polyunsaturated fatty acids
CS: Credible set
CUD: Cannabis use disorder
DHA: Docosahexaenoic acid
FADS: Fatty acid desaturase
GCKR: Glucokinase regulatory protein
GWAS: Genome-wide association study
h^2^: Heritability
INS: Insomnia
IVW: Inverse weighted variance
LA: Linoleic acid
LD: Linkage disequilibrium
LDSC: LD Score regression
MAF: Minor allele frequency
MDD: Major depression
MHC: Major histocompatibility complex
MOOD: Mood disorders
MR: Mendelian randomization
NAFLD: Nonalcoholic fatty liver disease
nc12: Number of common variants shared between trait1 and trait2
NE: Neuroticism
OCD: Obsessive-compulsive disorder
OD: Opioid dependence
PGC: Psychiatric Genomic Consortium
PP: Posterior probability
PTSD: Post-traumatic stress disorder
PUFA: Polyunsaturated fatty acids
r_g_: Genetic correlation coefficient
SCZ: Schizophrenia
SNP: Single-nucleotide polymorphism
TS: Tourette syndrome
VEP: Variant Effect Predictor

## Acknowledgements

Research reported in this publication was supported by the National Institute Of General Medical Sciences of the National Institutes of Health under Award Number R35GM143060. The content is solely the responsibility of the authors and does not necessarily represent the official views of the National Institutes of Health. We thank the PGC, IEU Open GWAS, and CNCR for providing access to genome-wide association study data. We also thank all research participants who provided DNA samples for these studies. We thank the UGA GACRC staff for facilitating our data analysis.

## Author contributions

K.Y. conceptualized and supervised the study. K.Y., H.X., and Y.S. designed the analysis. H.X. collected data. H.X. and Y.S. performed most data analysis with assistance from M.F., N.T.R.M., and C.F.C.. H.X., Y.S., K.Y., J.T.B., and C.W.K.C. interpreted the results and wrote the manuscript. All authors reviewed, revised, and approved the manuscript.

