## Supplementary Figures for "Shared genetic basis informs the roles of polyunsaturated fatty acids in brain disorders"

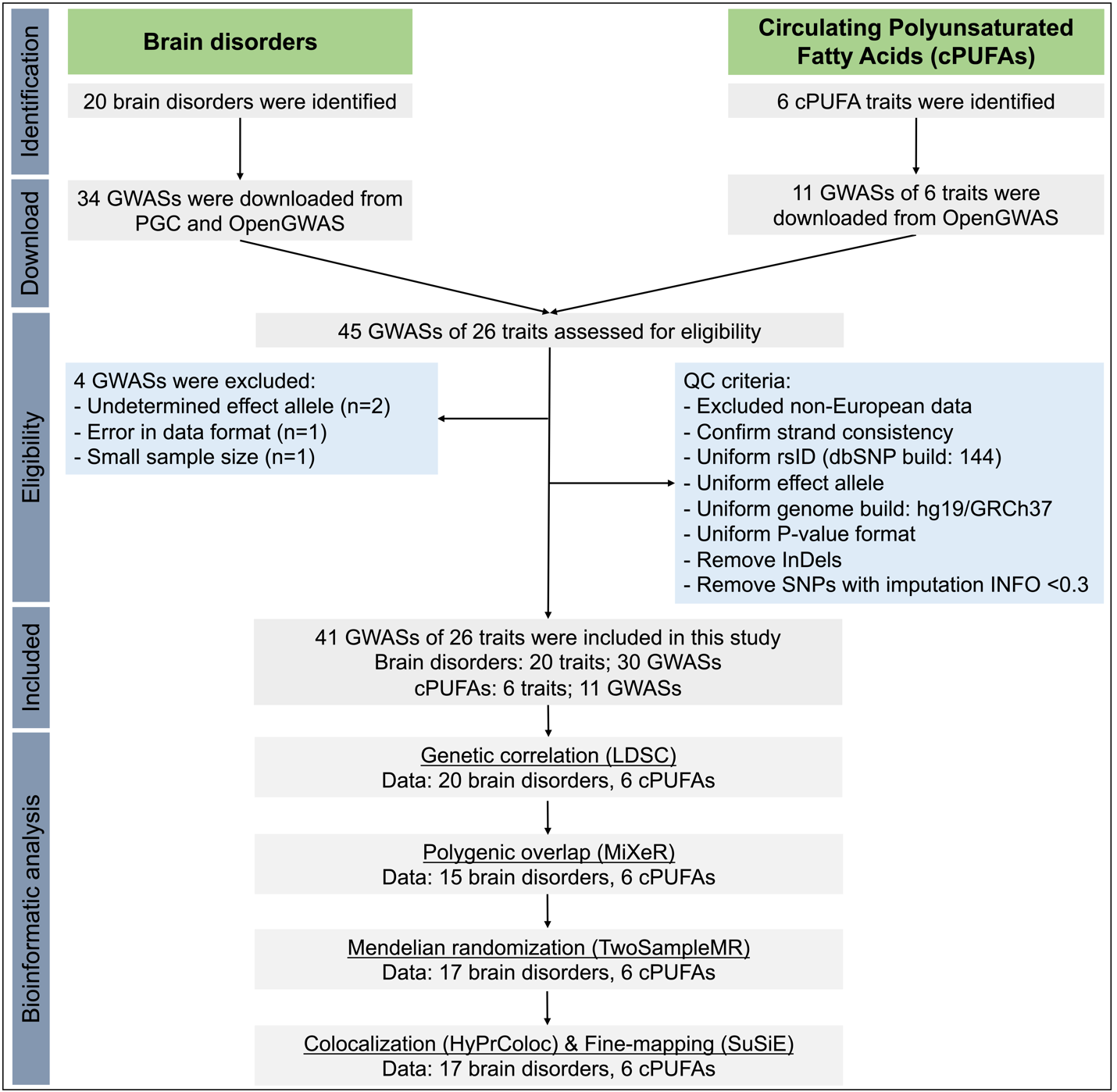

**Figure S1.** **Overview of the study.**

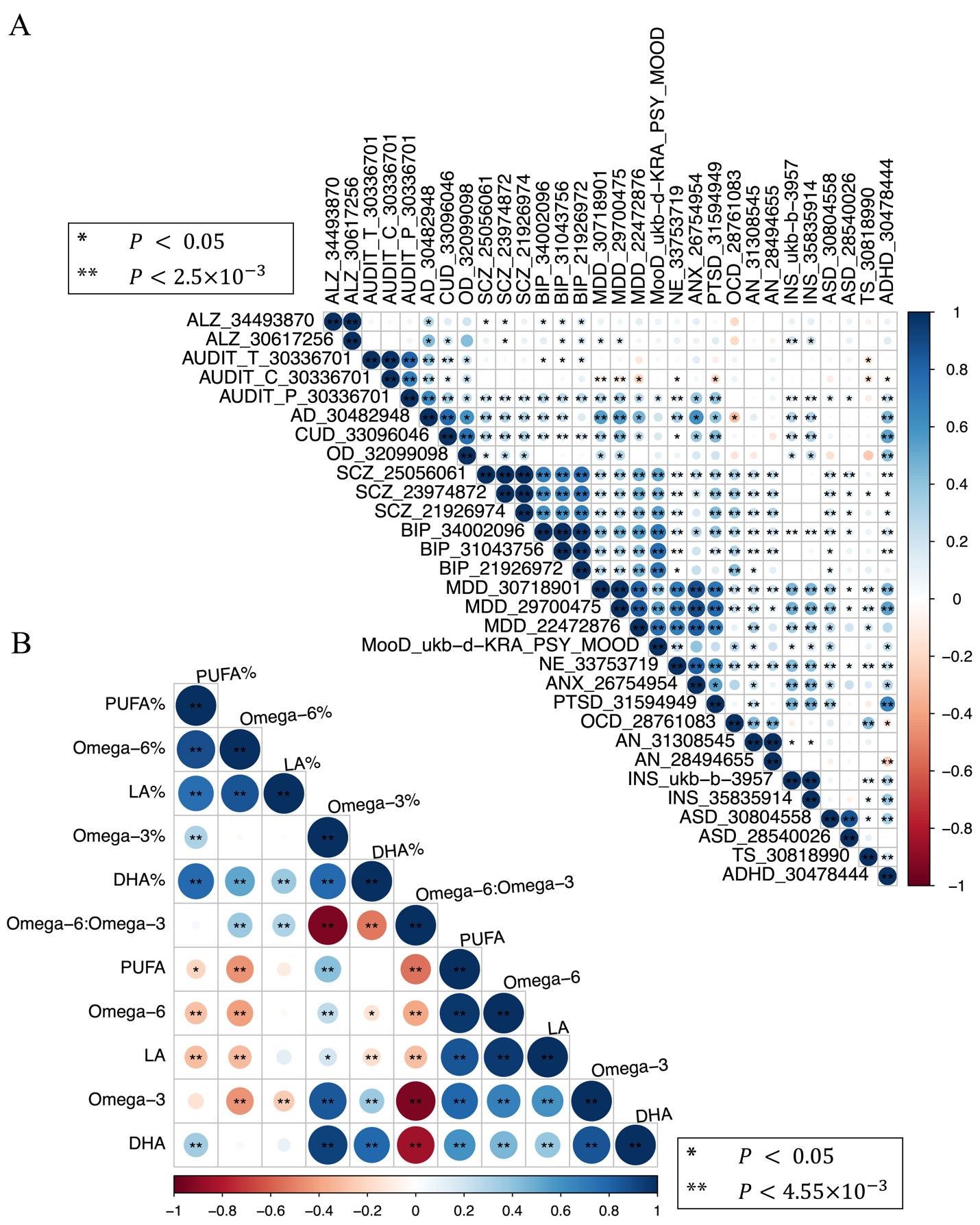

**Figure S2. Pairwise genetic correlations A) between brain disorders and B) between cPUFA phenotypes.** GWAS summary statistics of brain disorders are presented in a format with the phenotype_PMID. P-value cutoffs of 0.05, 0.05/# of tests were used to represent different levels of statistical significance; colors are used to represent degree of genetic correlation (r_g_) between two traits.

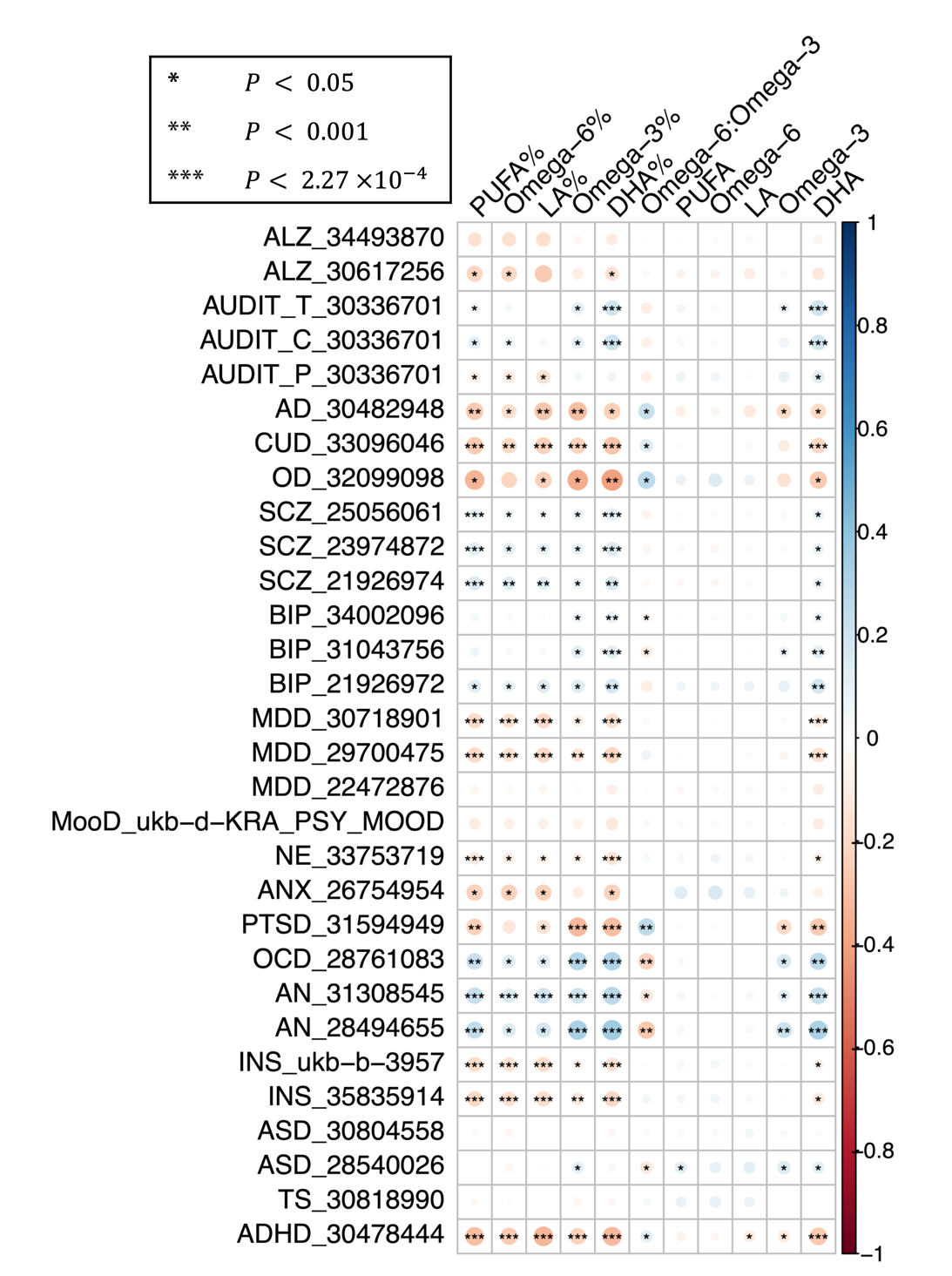

**Figure S3. Pairwise genetic correlations between cPUFAs and brain disorders.** P-value cutoffs of 0.05, 0.001, 4.17×10^-4^ were used to represent different levels of statistical significance; colors are used to represent degree of genetic correlation (r_g_) between two traits.

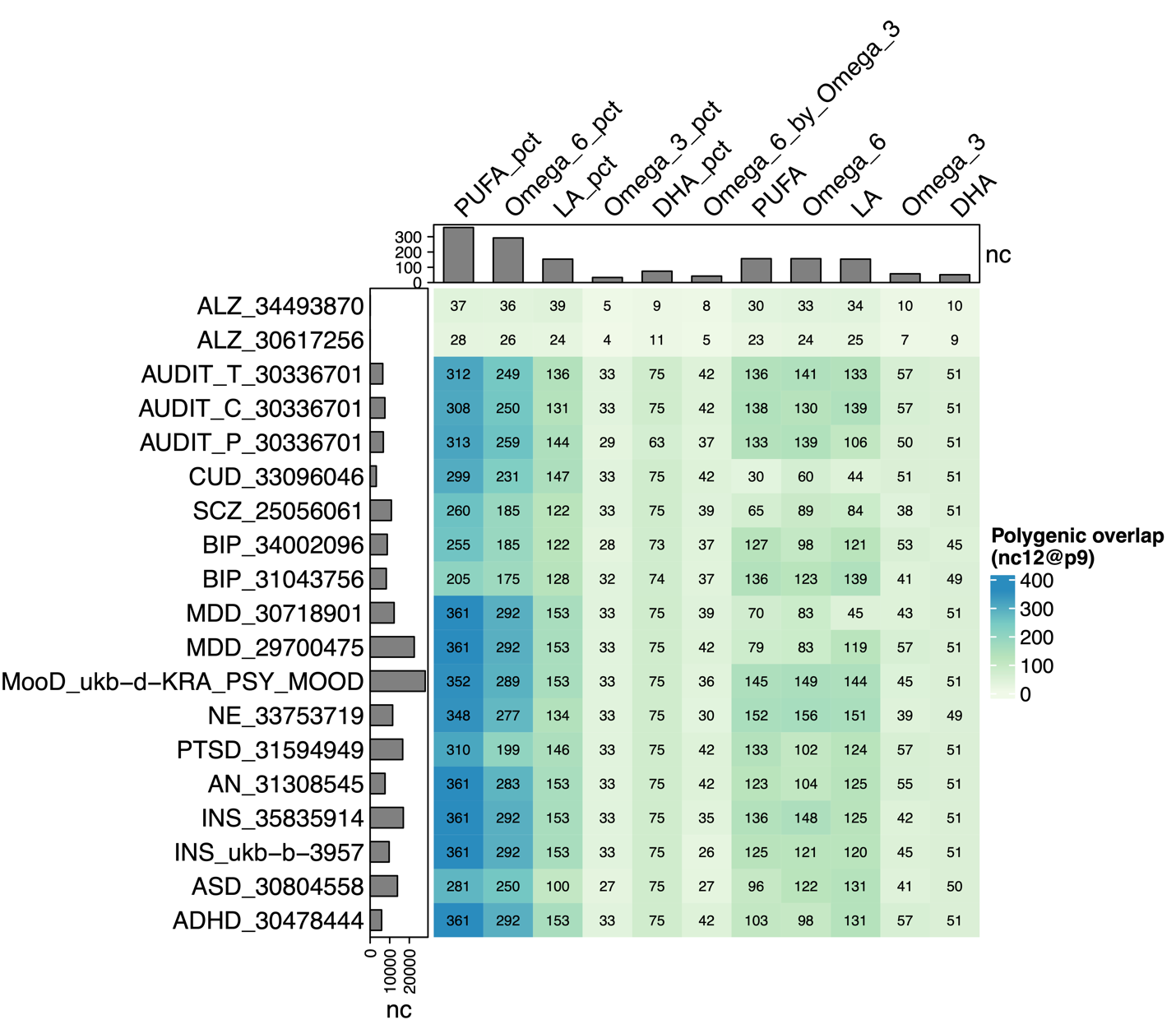

**Figure S4. Pairwise polygenic overlaps between cPUFAs and brain disorders.** The color and number of each box indicate the degree of polygenic overlap and number of causally associated SNPs shared between cPUFAs and brain disorders (nc_12_). Barplots on the top and left indicate the number of cPUFAs- and brain disorders- associated variants, respectively, which explain 90% of SNP-based heritability.

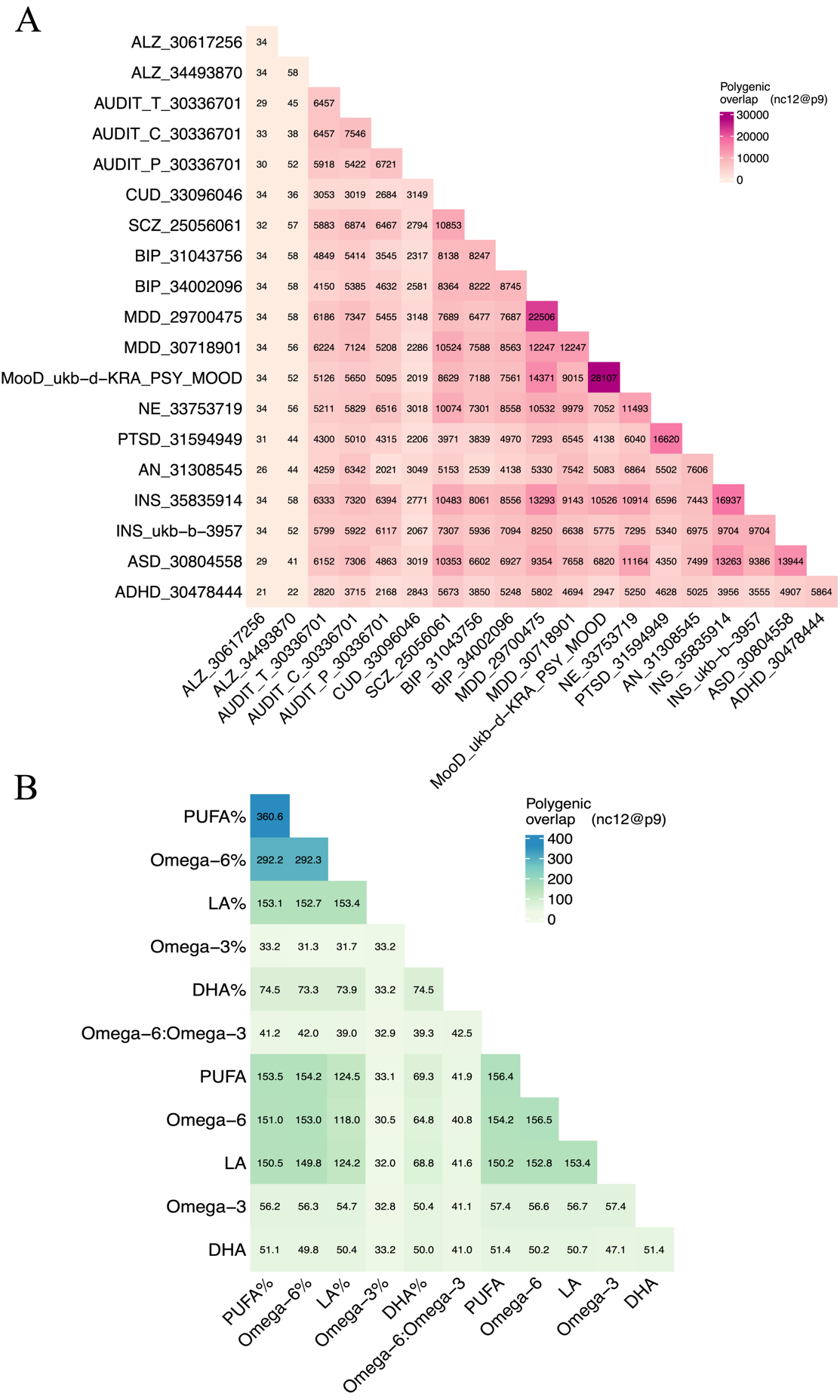

**Figure S5. Pairwise polygenic overlaps** **A) between brain disorders and B) between cPUFAs.** The color and number of each box indicate the degree of polygenic overlap and number of causally associated SNPs that explain 90% of SNP-based heritability, shared between cPUFAs or between brain disorders (nc_12_).

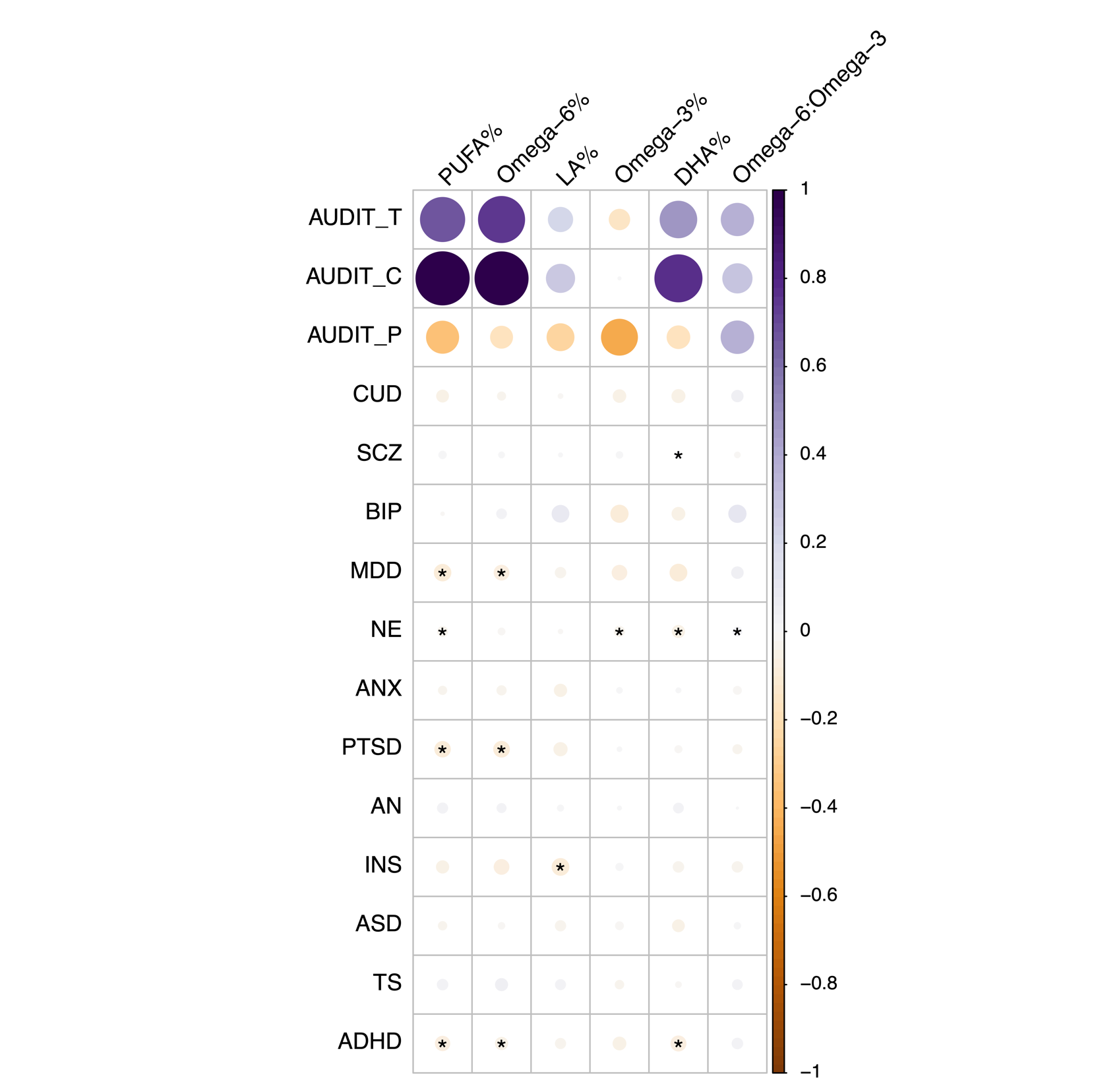

**Figure S6. Reverse MR results.** Heatmap summarized the causal effects of 15 brain disorders on 6 cPUFAs. Except for 4 brain disorders (ANX, ASD, PTSD, TS), IVW *P* value of 0.05 was used to represent statistical significance; colors are used to represent the causal effects (β_IVW_) of cPUFAs on brain disorders. Causal effect of four brain disorders (ANX, ASD, PTSD, TS) on cPUFAs and corresponding *P* values were estimated using Wald ratio test.

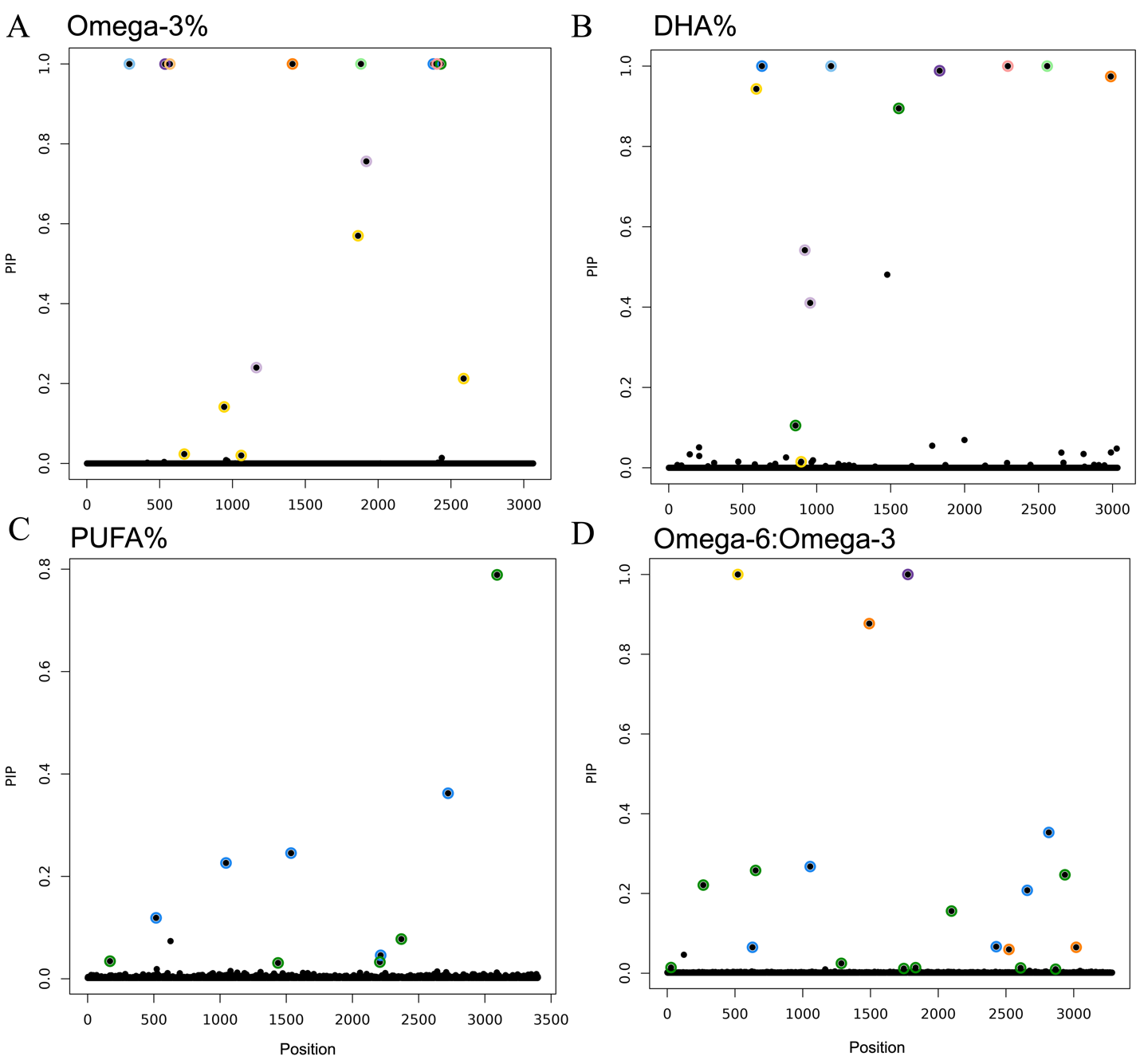

**Figure S7. Statistical fine-mapping posterior inclusion probability plots in 11q12.1-11q12.3 for A) omega-3%, B) DHA%, C) PUFA% and D) omega-6:omega-3.** Variant position is shown on x axis, posterior inclusion probability (PIP) on y axis. Colors are used to represent different 95% credible sets. Each 95% credible set contains a causal variant. There are multiple putative causal variants in the region for omega-3%, DHA%, PUFA% and omega-6:omega-3.

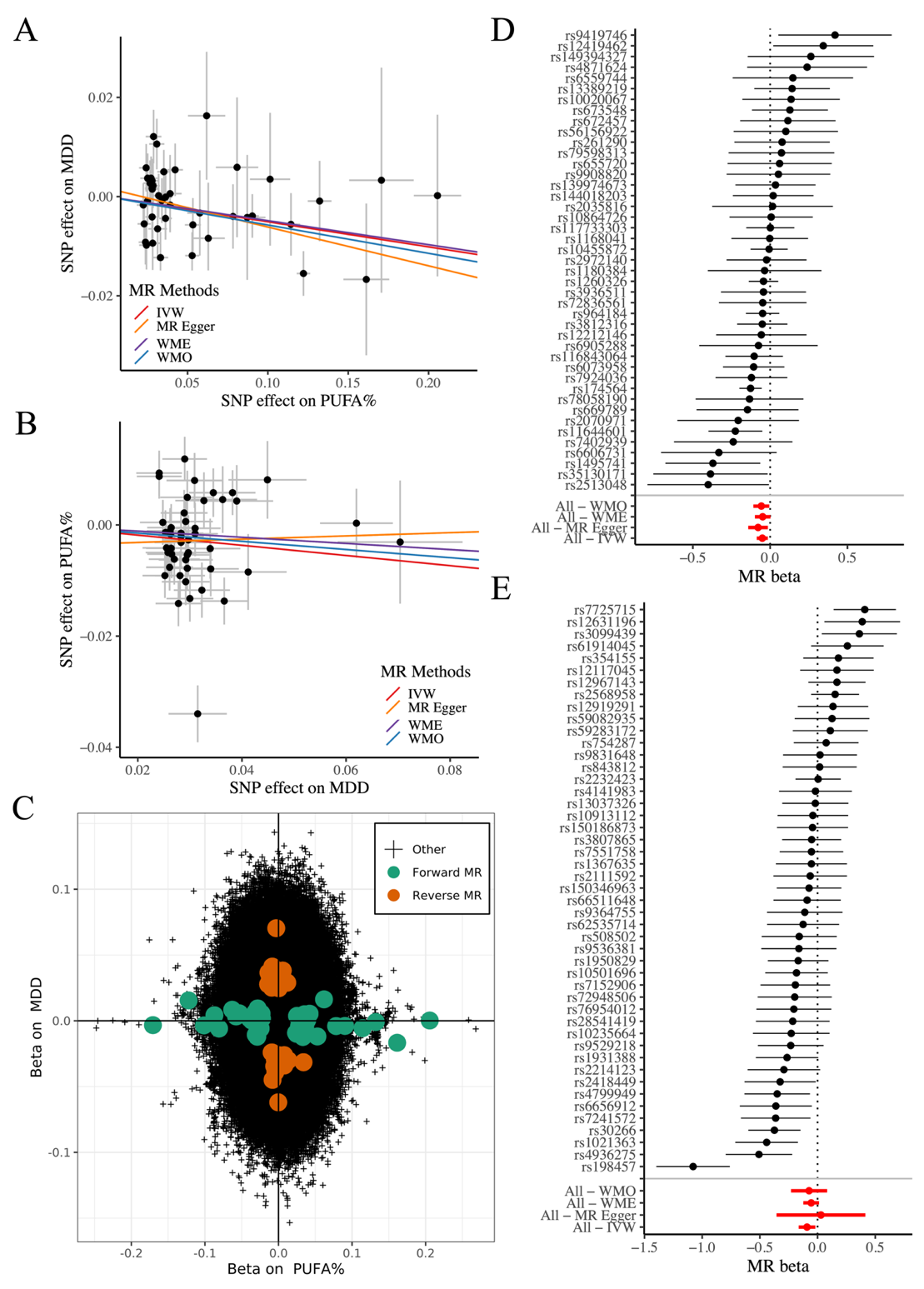

**Figure S8. Bidirectional association between PUFA% and MDD. A)** Forward MR estimated causal effects of PUFA% (x axis) on MDD (y axis). Causal effect estimated by the four models are shown by fitted lines; slopes of these lines indicate causal effect sizes. **B)** Reverse MR estimated causal effects of MDD (x axis) on PUFA% (y axis). **C)** Genetic effect of genome-wide SNPs on PUFA% (x axis) and MDD. Genetic variants that are selected in forward and reverse MR analysis are shown in green and red, respectively. **D)** Forest plot shows 43 genetic instruments included in the PUFA%-MDD forward MR analysis. Causal effect estimated by the four models are marked in red. **E)** Forest plot shows 48 genetic instruments included in the MDD-PUFA% reverse MR analysis.

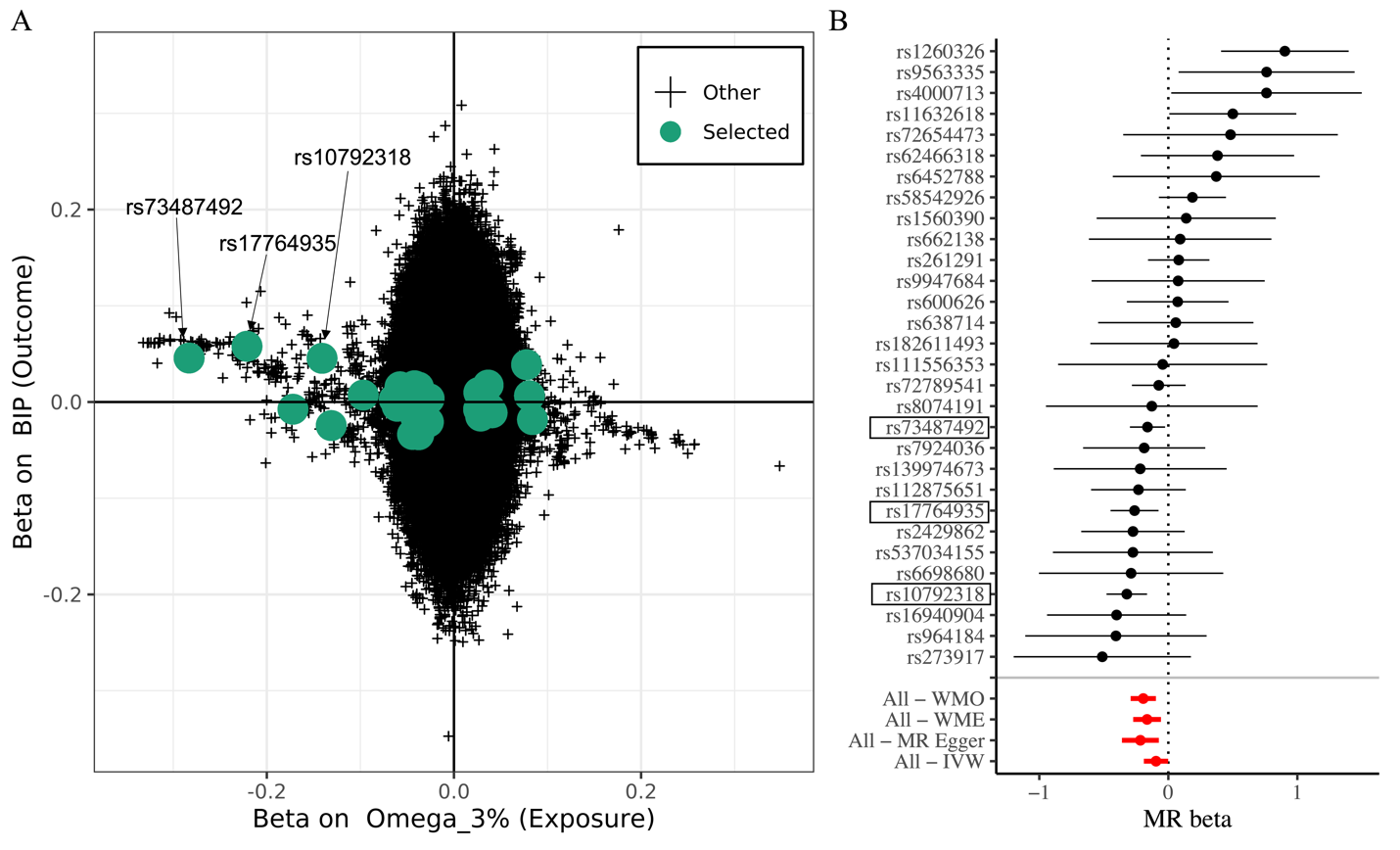

**Figure S9.** **Differences in the SNP sets in omega-3%-BIP genetic correlation and MR analysis. A)** Genetic effect of genome-wide SNPs included in genetic correlation analysis on omega-3% (x axis) and BIP (y axis). Genetic variants that are selected in omega-3%-BIP MR analysis are shown in green. Three highlighted SNPs are in the FADS locus. **B)** Forest plot shows 30 genetic instruments included in the omega-3%-BIP MR analysis. Causal effect estimated by the four models are marked in red.
